# Serological and Molecular Study on Caprine Brucellosis in Puducherry (India) and its Public Health Significance

**DOI:** 10.1101/2022.06.24.22276885

**Authors:** Abhishek Madan, Gururaj Kumaresan, Bhanu Rekha V, Dimple Andani, Anil Kumar Mishra, Ajay Kumar VJ, Thanislass Jacob, Kavita Vasudevan P

**Author notes:** Corresponding author: Gururaj Kumaresan, Senior Scientist (Veterinary Microbiology), Indian Council of Agricultural Research – Central Institute for Research on Goats (ICAR – CIRG), Makhdoom, Farah (post), Mathura 281122, Uttar Pradesh, India. E mail.

## Abstract

Caprine brucellosis due to *Brucella melitensis* is an important zoonotic disease. The present study was carried out to address the lack of a comprehensive study on the status of caprine brucellosis in Puducherry, India using serological and molecular tests in goats and to assess the seroprevalence in human risk groups of the aforementioned region to ascertain the public health significance of the disease. Seroprevalence in 120 goats was found to be zero, 3.33% and 18.33% by Rose Bengal agglutination Test (RBT), Standard Tube Agglutination Test (STAT) and Immunoglobulin G Indirect Enzyme Linked Immune Sorbant Assay (IgG iELISA) respectively. Of the 120 goat genital swabs screened, while conventional polymerase chain reaction (PCR) detected genus specific *16S rRNA* and *Brucella melitensis* specific *omp2* genes in 17.50% and 5.00% of samples respectively, the *OMP31*TaqMan^®^ real time PCR with a positive detection of 40.0 % was both the most sensitive and specific for detection of *Brucella melitensis*. The study provides insight into the optimization of diagnostic tests following cluster wise sampling for brucellosis in goats. The strain of *Brucella melitensis* in Puducherry was found to be Biovar 3 based upon suggestive results of Restriction Fragment Length Polymorphism (RFLP) of *omp2* gene product. Seroprevalence by IgG iELISA was 33.33 % in 30 samples from human subjects. Serological evidence of caprine brucellosis in goats and human subjects and molecular detection of *Brucella melitensis* in Puducherry, India warrants regular screening, surveillance and reporting of disease in goats and human risk groups.

**IMPORTANCE:**

- Brucellosis is an important zoonotic pathogen causing abortions in domestic animals as well as posing risk to livestock keepers and handlers
- Control of this disease requires proper surveillance programme, and in this study the peninsular coastal region of India was sampled by a cluster method and evaluated for brucellosis in goats as well as people at risk
- This study compared the diagnostic sensitivity of various serological and molecular tests and found that the OMP31 gene TaqMan probe based Real Time assay to be highly sensitive in diagnosing brucellosis
- Similarly, in human subjects exposed to risk factors like animal handlers, veterinarians, livestock keepers the sero-positivity was 33.33% by ELISA

## INTRODUCTION

Caprine brucellosis due to *Brucella melitensis* (*B. melitensis*) is an important zoonotic disease. It is more prevalent in the tropical and sub-tropical regions of the world and is marked as a neglected tropical disease (NTD) with significant economic impact in developing countries such as India (1). The disease manifests itself in the goat populations primarily in the reproductive and placental tissues leading to abortion in female goats while generalized bacteraemia affecting a wide range of organ systems is also possible. The most virulent cause of brucellosis in human subjects is *B. melitensis* (2, 3). Human risk groups such as veterinarians, farmers, laboratory personnel etc. may contract infection from goat populations and show wide range of clinical symptoms affecting various organ systems of which Undulating Fever (Pyrexia of Unknown Origin) is the most commonly manifested (4).

Although isolation by culture is the gold standard for diagnosis of brucellosis, the zoonotic nature of the bacteria, long incubation conditions and need for bio-safety cabinets form considerable drawbacks (5, 6). Thus, various indirect (serological) and direct (molecular) tests are envisaged for the screening and diagnosis of brucellosis in both goats as well as human subjects. Of the indirect tests, the Rose Bengal agglutination Test (RBT), Standard tube agglutination test (STAT), 2 Mercaptoethanol precipitation test (2ME) and Enzyme Linked Immune Sorbant Assays (ELISA’s) are used predominantly for knowing the status of brucellosis in goat populations (7) as well as humans due to *B. melitensis* (2). As far as molecular screening is considered, various types of polymerase chain reactions (PCR) such as simple or multiplex, real time systems (SYBR^®^Green or TaqMan^®^ probes) and Loop mediated isothermal Amplification (LAMP) have been used which target genes either at the genus level (*bscp31, IS1711* or *16S rRNA*) or species level (*omp31, omp2,* and genes coding for other outer membrane proteins) for *B. melitensis* (7, 8). For strain identification, Restriction Fragment Length Polymorphisms (RLFP) of various gene products (*omp22, omp25, omp31* and *omp2*) has been used. Other methods include sequencing using Multiple Locus Variable tandem Analysis (MLVA) or Multi Locus Sequencing analysis (MLSA) (9).

Studies on brucellosis in goats in various states of India show that the seroprevalence ranges from 1.14 % to 18.77 % (10, 11). Also, molecular screening from the suspected field samples has revealed that *B. melitensis* DNA can have an occurrence between 13.33 % and 49.05 % (12, 13) based on nucleic acid amplification tests. Molecular typing suggests that the most predominant variant of the *B. melitensis* is Biovar 3 in the country of India (12) while strain 16M has also been reported (14). In at-risk human subjects, various studies on brucellosis in India show the seroprevalence in the range of 1.60 % to 16.52 % (15, 16).

Puducherry region of the Union Territory (UT) of Puducherry, India has a population of 27,623 goats (17). Although prior work on caprine brucellosis has been carried out in various regions of India, to address the lack of a comprehensive study on caprine brucellosis due to *B. melitensis* in Puducherry, India and its public health significance in human risk groups, the present study was carried out with the following objectives: First, to report and analyse the status of caprine brucellosis due to *B. melitensis* based on serological tests and nucleic acid amplification techniques following scientific cluster sampling of the goat populations in Puducherry and second, to know the seroprevalence of brucellosis in human risk groups of Puducherry, India to ascertain the zoonotic implications of the disease.

## MATERIALS AND METHODS

### Ethical and regulatory approval

The study was approved by the Institute Research Committee at Rajiv Gandhi Institute of Veterinary Education and Research (RIVER), Puducherry, India. For human sampling, ethical permission and approval was obtained from Institute Ethics Committee (IEC), Indira Gandhi Medical College and Research Institute (IGMC & RI), Puducherry, India, vide letter No. 305/IEC-30/IGMC&RI/PP/2020 dated 02.12.2020.

### Sampling plan

For sampling of goats in the study, 12 clusters (3 farms/organized clusters and 9 villages/unorganized clusters) were identified randomly in various regions of Puducherry in UT of Puducherry based on National Animal Disease Referral Expert System (NADRES) sampling tool developed at Indian Council of Agricultural Research - National Institute of Veterinary Epidemiology and Disease Informatics (ICAR-NIVEDI), Bengaluru, India. The sampling regions covered all major communes and geographically variable zones (Supplement I) The sampling of goats was carried out during late September to early November of the calendar year 2020. The average elevation of the sampling areas was 10 metres above mean sea level. The climatological parameters included average high temperatures of 33.1°C, 31.5°C and 29.8°C and average low temperatures of 24.9°C, 24.5°C and 23.6°C for the months of September, October and November respectively. The average rainfall in millimetres (mm) was usually 132.8, 273.9 and 350 for the months of September, October and November respectively (18). Blood (for serum) and genital swabs were collected from 10 goats randomly in each cluster (total of 120) comprising both adult male and female animals above 6 months of age.

Thirty (30) blood samples were collected from identified willing human risk groups such as veterinary interns, post graduate students and farmers (under the supervision of medical professional) over 18 years of age from Puducherry, India along with pertinent details such as their age, gender, history of animal handling and other co-morbidities (if any).

### Collection and processing of samples

#### Goat blood and genital swabs

From each goat, approximately 6 millilitre (ml) of blood was collected by jugular venepuncture following sterile/aseptic precautions. After clot retraction, the serum obtained was transferred to microfuge tubes and centrifuged at 1200 rpm (rotations per minute) for 7 minutes.

Genital swabs were taken from both male and female animals using sterile screw cap swabs (Hi-Media^TM^) giving a contact time of 10 to 20 seconds on the genital mucous membrane. The swabs were reconstituted with 1 ml of sterile phosphate buffered saline (PBS) (pH 7.2 to 7.4) and vortexed for 5 minutes.

#### Human blood samples

Blood (approximately 4 ml) was collected using the standard protocol approved by the Indian Council of Medical Research (ICMR) after obtaining written consent from the participants.

All samples (blood serum or reconstituted swabs) obtained were maintained either at 4°C (short term storage) or minus 20°C (long term storage) in sterile cryo-tubes until use.

### Serological tests on goat blood serum

#### RBT

The Rose Bengal Brucella coloured antigen was procured from Institute of Animal Health and Veterinary Biologicals (IAHVB), Bengaluru, India and samples were tested using standard procedure described by Alton et al. (19) and accordingly graded (+++, ++, + or Negative) based on the degree of agglutination.

#### STAT

The sera samples were tested as per the protocol outlined by OIE (20) with slight modifications (serum was serially double diluted using carbol saline instead of normal saline) using the *Brucella melitensis* killed antigen (produced in-house at ICAR-CIRG according to OIE protocols). Following incubation at 37°C for 48 to 96 hours following the tubes were read either as positive result based on mesh/lattice formation or negative based on pellet formation. Positive and negative controls were maintained for comparison and interpretation of the results. For goat serum, agglutination at 1:40 or above dilutions was taken as positive by STAT.

#### 2ME

Serum samples from goats positive in STAT were subjected to 2ME test as per the standard protocol (20) using killed antigen produced in-house at ICAR-CIRG. The reading of the 2ME test was compared with the reading of STAT for goat serum samples and accordingly interpretation was done to decipher the stage of infection.

#### IgG iELISA

The procedure followed for IgG iELISA was standardized and developed at microbiology laboratory of ICAR-CIRG using *B. melitensis* protoplasmic antigen (BruLISA^®^ Kit) (13,21,22). Absorbance was read at 450 nm (nanometres) in ELISA reader (iMark^TM^ Microplate Reader, BioRad^®^) and the blanked values of the OD (Optical Density) were used to calculate the sample to positive ratio (S/P ratio) which was then subjected to logical analysis.

### Molecular screening of goat genital swabs

From the reconstituted swab samples, DNA was extracted by using QIAmp^TM^ DNA Mini Kit (#51304, Qiagen^®^, USA) by following the manufacturer’s protocol. For molecular screening, the extracted DNA samples were divided into 5 different super-pool sets (n=24 sample cocktail DNA for each super-pool for swab DNA). Only positive super pools were divided into mini-pools (n=6). If any mini-pool showed positivity, all the individual samples in that mini-pool were subjected to PCR/qPCR. Positive control (PC) and No template control /negative control (NC/NTC) were used at every hierarchy of the samples tested.

### Conventional PCR

Conventional PCR was carried out for brucella genus specific *16S rRNA* gene and *B. melitensis* specific genes (*omp31* and *omp2*) (Table 1) by the use of standardized PCR conditions (Supplement II). The PCR products thus obtained were subjected to 1.8 % submarine agarose gel electrophoresis at 100 V for 40 minutes. The gel was then visualized under ultra violet transilluminator (Alpha Innotech) following which the images were documented and interpreted as positive or negative based on the appearance of the band or the absence of the specific product band respectively.

**TABLE 1.**
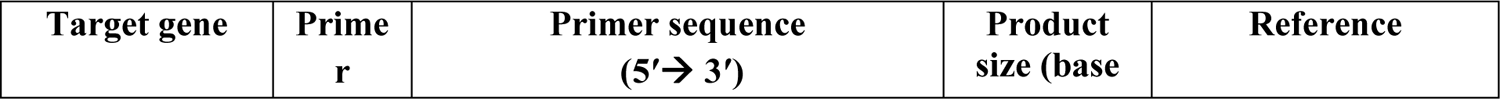
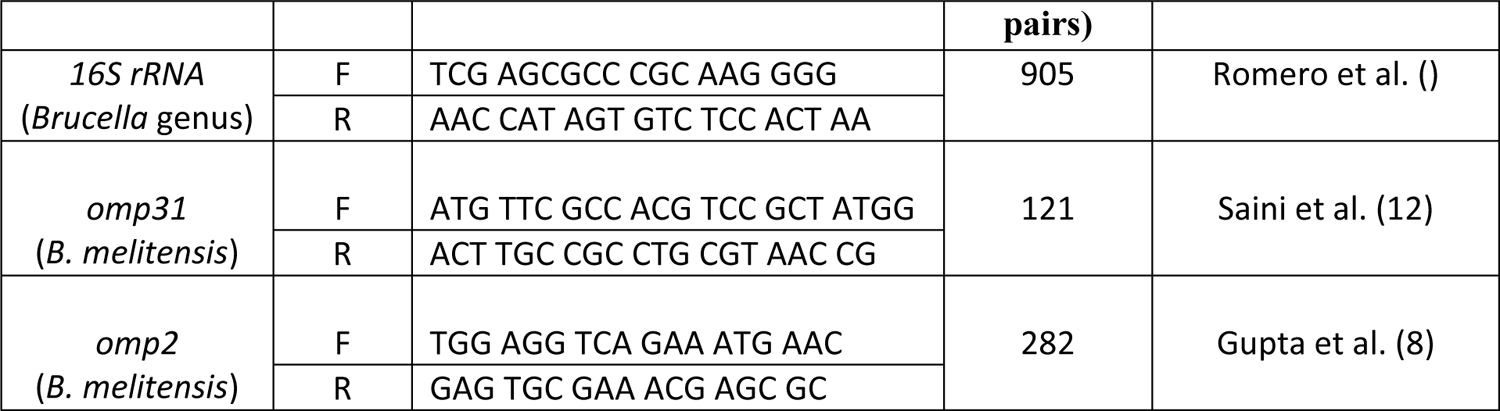
List of genes targeted by conventional PCR for *Brucella*

### OMP31 TaqMan^®^ probe based real time PCR

A TaqMan^®^ probe based real-time PCR for the analysis of *omp31* gene (open reading frame) of *B. melitensis* was developed and standardized at the microbiology laboratory of ICAR-CIRG (13). The reaction was carried out in CFX96™ Real-time system (Bio-Rad^®^, USA) by using the primer set (Forward: 5’ATG TTC GCC ACG TCC GCT ATGG 3’; Reverse: 5’ACT TGC CGC CTG CGT AAC CG 3’) and probe (6-FAM-5’TCCTGTTGACACCTTCTCGTGG-3’-BHQ-1) under standardized conditions (Supplement II). The Relative Fluorescence Units (RFU) at the beginning and the end of the reaction mixture in concurrence with the Cq value (no. of cycles wherein sample curve intersects threshold line) was obtained following which the average RFU was found out. Sample average RFU of over and above 50 per cent of the average RFU of positive control in the reaction set and based upon logical analysis was considered to be positive whereas an average RFU over and above 100 per cent of the positive control’s average RFU was taken as strongly positive for the *B. melitensis* DNA in the samples tested.

### Analyses of battery of tests for caprine brucellosis

The results of the tests *viz*. RBT, STAT, IgG iELISA, *16S rRNA* PCR, *omp31* PCR and *omp 2* PCR were compared for their Sensitivity, Specificity, Diagnostic accuracy, Positive predictive value (PPV) and Negative predictive value (NPV) with respect to *OMP31* TaqMan^®^ qPCR (23). The tests were also analyzed in pairs for their goodness of fit by using Kappa Value (24) and McNemar’s test (25).

### PCR - RFLP

Samples positive for *omp2* gene by conventional PCR were subjected to RFLP using the *Pst*I-HF restriction enzyme. The reaction mixture was incubated at 37°C for 30 minutes and followed by inactivation of enzyme at 65°C for 5 minutes and then maintained at 4°C in Veriti^TM^ 96 well thermal cycler (Life Technologies, USA) until loaded into the gel. Digested *omp2* PCR product and undigested *omp2* PCR product (10 μl) were added along with DNA markers. Electrophoresis was then carried out at constant voltage of 80 V for 75 minutes. Then, the gel was visualized under UV trans-illuminator and images were documented and interpreted accordingly (12).

### Serological tests on human blood serum samples

Blood serum samples from human subjects were tested for brucellosis using RBT, STAT and IgG iELISA. While the procedure followed for RBT and STAT was similar to that of goat samples; STAT agglutination at dilutions over and above 1:160 were considered positive for human subjects (2). In the case of IgG iELISA, the protocol previously mentioned (21, 22) was used with the modification that rabbit anti-human IgG was used as secondary antibody.

### Statistical analysis

The categorical data were expressed as percentage and the significance of differences was checked by Chi-squared test (χ^2^ Test) using GraphPad^®^ Prism (v 5.0) software at 95 % confidence interval (α = 0.05).

## RESULTS AND DICSUSSION

In total, 120 samples (set of blood and genital swab from each animal) were collected from 3 organized and 9 unorganized clusters in Puducherry, India. The category wise details of the samples revealed that 20 animals were males, 100 were females. Also, it was seen that 63, 54 and 3 animals belonged to the up to 1 year, 1 to 2 years and above 2 years age groups, respectively. Thirty animals belonged to the organized group of clusters while 90 belonged to the unorganized group (Table 2).

**TABLE 2.** Tabulation of category wise results of battery of tests for Caprine Brucellosis (RBT, STAT, IgG iELISA, *16S rRNA* PCR, *omp31* PCR, *omp 2* PCR and *OMP31* TaqMan^®^ qPCR) and their per cent positivity in various clusters of Puducherry

| Category | T | RBT |  | STAT |  | IgG iELISA |  | Conventional PCR |  |  |  |  |  | OMP31TaqMan®<br>qPCR |  |
| --- | --- | --- | --- | --- | --- | --- | --- | --- | --- | --- | --- | --- | --- | --- | --- |
|  |  |  |  |  |  |  |  | 16S rRNA |  | omp31 |  | omp2 |  |  |  |
|  |  | P | % | P | % | P | % | P | % | P | % | P | % | P | % |
| Sex wise |  |  |  |  |  |  |  |  |  |  |  |  |  |  |  |
| Male | 20 | 0 | 0.00 | 0 | 0.00 | 1 | 5.00 | 3 | 15.00 | 0 | 0.00 | 0 | 0.00 | 8 | 40.00 |
| Female | 100 | 0 | 0.00 | 4 | 4.00 | 21 | 21.00 | 18 | 18.00 | 0 | 0.00 | 6 | 6.00 | 40 | 40.00 |
| χ² Test |  | NA |  | NA |  | NA |  | NA |  | NA |  | NA |  | 2.963<sup>NS</sup> (p=0.0851) |  |
| Age group wise |  |  |  |  |  |  |  |  |  |  |  |  |  |  |  |
| Up to 1 yr. | 63 | 0 | 0.00 | 1 | 1.59 | 11 | 17.46 | 11 | 17.46 | 0 | 0.00 | 4 | 6.35 | 20 | 31.75 |
| 1 to 2 years | 54 | 0 | 0.00 | 3 | 5.56 | 11 | 20.37 | 8 | 14.81 | 0 | 0.00 | 2 | 3.70 | 26 | 48.15 |
| Above 2 yrs. | 3 | 0 | 0.00 | 0 | 0.00 | 0 | 0.00 | 2 | 66.67 | 0 | 0.00 | 0 | 0.00 | 2 | 66.67 |
| χ² Test |  | NA |  | NA |  | NA |  | NA |  | NA |  | NA |  | NA |  |
| Major Clusters wise |  |  |  |  |  |  |  |  |  |  |  |  |  |  |  |
| Organized | 30 | 0 | 0.00 | 2 | 6.67 | 8 | 26.67 | 0 | 0.00 | 0 | 0.00 | 0 | 0.00 | 3 | 10.00 |
| Unorganized | 90 | 0 | 0.00 | 2 | 2.22 | 14 | 15.56 | 21 | 23.33 | 0 | 0.00 | 6 | 6.67 | 45 | 50.00 |
| χ² Test |  | NA |  | NA |  | 1.855<sup>NS</sup> (p=0.1731) |  | NA |  | NA |  | NA |  | NA |  |

T – No. tested
P – No. positive
% - Per cent positive
NA – Not applicable
NS – Not significant

### Serological tests on goat blood serum

The seroprevalence of brucellosis in 120 goat blood serum samples by RBT, STAT and IgG iELISA was zero, 3.33 % and 18.33 %, respectively (Table 3). One of the plate systems of IgG iELISA on goat serum was also shown (Fig. 1). The split up of the results of serological tests in goat serum samples based on the sex of the animal/s, age group and also the major clusters was accordingly tabulated previously (Table 2).

**FIG 1.**
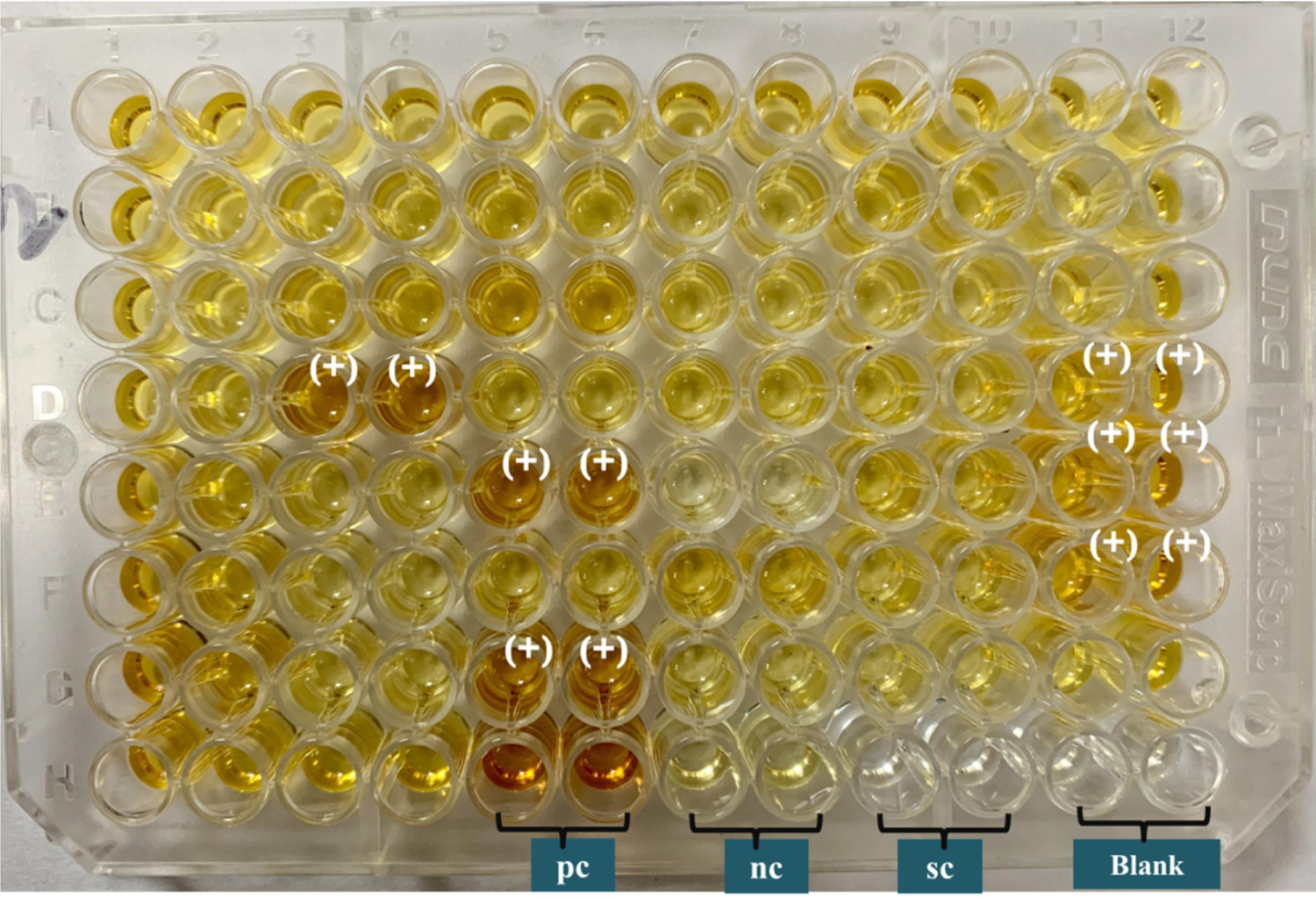
Plate depicting IgG iELISA in goat blood serum: (+) Positive samples, pc (Positive Control), nc (Negative Control), sc (Substrate control) and Blank

**TABLE 3.**
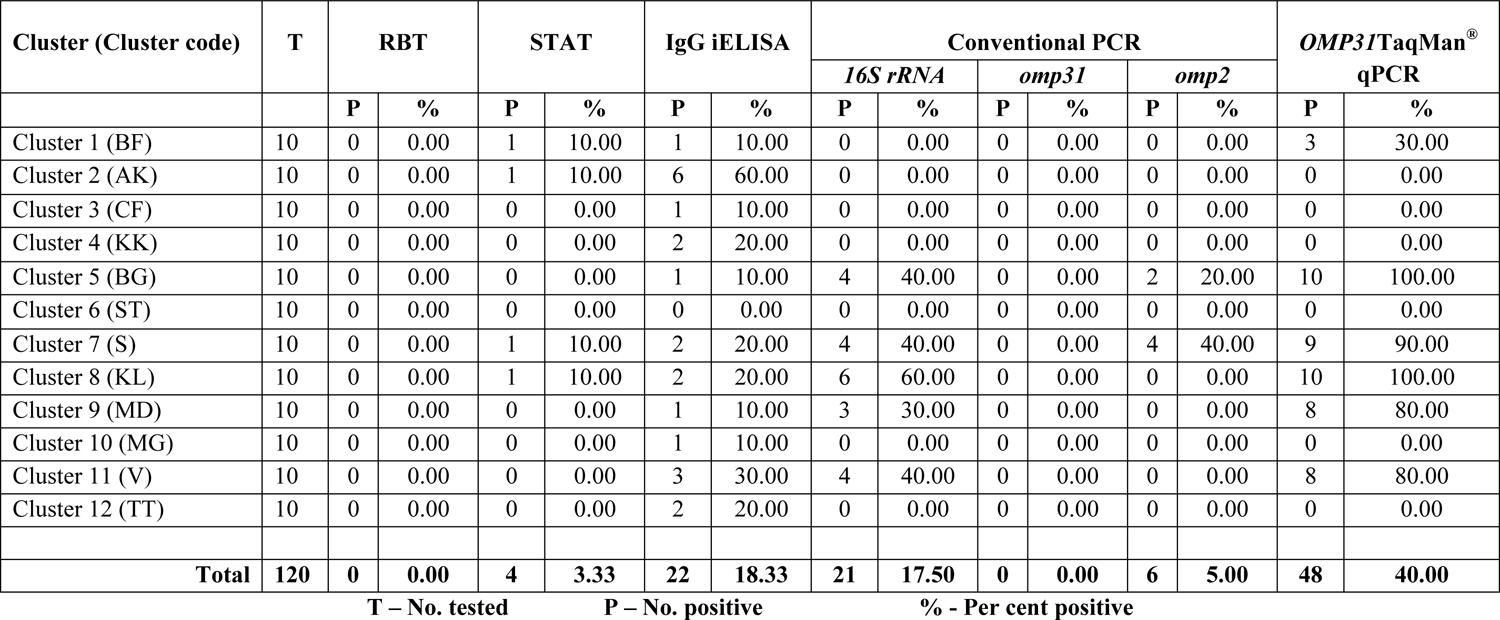
Tabulation of results of battery of tests for Caprine Brucellosis (RBT, STAT, IgG iELISA, *16S rRNA* PCR, *omp31* PCR, *omp 2* PCR and *OMP31* TaqMan^®^ qPCR) and their per cent positivity in various clusters of Puducherry

| Cluster (Cluster code) | T | RBT |  | STAT |  | IgG iELISA |  | Conventional PCR |  |  |  |  |  | OMP31/TaqMan<sup>®</sup> qPCR |  |
| --- | --- | --- | --- | --- | --- | --- | --- | --- | --- | --- | --- | --- | --- | --- | --- |
|  |  | P | % | P | % | P | % | *16S rRNA* |  | *omp31* |  | *omp2* |  | P | % |
| Cluster 1 (BF) | 10 | 0 | 0.00 | 1 | 10.00 | 1 | 10.00 | 0 | 0.00 | 0 | 0.00 | 0 | 0.00 | 3 | 30.00 |
| Cluster 2 (AK) | 10 | 0 | 0.00 | 1 | 10.00 | 6 | 60.00 | 0 | 0.00 | 0 | 0.00 | 0 | 0.00 | 0 | 0.00 |
| Cluster 3 (CF) | 10 | 0 | 0.00 | 0 | 0.00 | 1 | 10.00 | 0 | 0.00 | 0 | 0.00 | 0 | 0.00 | 0 | 0.00 |
| Cluster 4 (KK) | 10 | 0 | 0.00 | 0 | 0.00 | 2 | 20.00 | 0 | 0.00 | 0 | 0.00 | 0 | 0.00 | 0 | 0.00 |
| Cluster 5 (BG) | 10 | 0 | 0.00 | 0 | 0.00 | 1 | 10.00 | 4 | 40.00 | 0 | 0.00 | 2 | 20.00 | 10 | 100.00 |
| Cluster 6 (ST) | 10 | 0 | 0.00 | 0 | 0.00 | 0 | 0.00 | 0 | 0.00 | 0 | 0.00 | 0 | 0.00 | 0 | 0.00 |
| Cluster 7 (S) | 10 | 0 | 0.00 | 1 | 10.00 | 2 | 20.00 | 4 | 40.00 | 0 | 0.00 | 4 | 40.00 | 9 | 90.00 |
| Cluster 8 (KL) | 10 | 0 | 0.00 | 1 | 10.00 | 2 | 20.00 | 6 | 60.00 | 0 | 0.00 | 0 | 0.00 | 10 | 100.00 |
| Cluster 9 (MD) | 10 | 0 | 0.00 | 0 | 0.00 | 1 | 10.00 | 3 | 30.00 | 0 | 0.00 | 0 | 0.00 | 8 | 80.00 |
| Cluster 10 (MG) | 10 | 0 | 0.00 | 0 | 0.00 | 1 | 10.00 | 0 | 0.00 | 0 | 0.00 | 0 | 0.00 | 0 | 0.00 |
| Cluster 11 (V) | 10 | 0 | 0.00 | 0 | 0.00 | 3 | 30.00 | 4 | 40.00 | 0 | 0.00 | 0 | 0.00 | 8 | 80.00 |
| Cluster 12 (TT) | 10 | 0 | 0.00 | 0 | 0.00 | 2 | 20.00 | 0 | 0.00 | 0 | 0.00 | 0 | 0.00 | 0 | 0.00 |
| **Total** | **120** | **0** | **0.00** | **4** | **3.33** | **22** | **18.33** | **21** | **17.50** | **0** | **0.00** | **6** | **5.00** | **48** | **40.00** |

T – No. tested
P – No. positive
% - Per cent positive

**All samples from goats were tested negative by RBT** which was in conflict to other reports in India which have reported a seroprevalence ranging from 1.42 % to 20.83 % (10, 26–29). The reasons for the negative RBT in the present study may be the following: First, the coloured antigen used in the current study was a *Brucella abortus* antigen (as is the one available for field testing conditions). Though cross reactions may occur due to low specificity of RBT, it is possible for the test to miss out on noticeable agglutination leading to false negatives (30) by virtue of the physiological properties of the goat serum (2). Second, there may be the absence of sufficient titre of antibodies. To overcome this, a protocol recommended by the OIE (20) which uses 25μl of the coloured antigen and 75μl of the blood serum may be explored. The present study projects the need for development of standardized *B. melitensis* coloured antigen/s specifically for serological diagnosis of brucellosis in small ruminants, especially goats for tests such as RBT.

**While the seroprevalence by STAT in goats** in the present study falls in similar range to a study in Jammu, India i.e. 3.42 % (31), it was below other reports from various parts of India which showed seropositivity ranging from 7.96 per cent to 17.68 per cent (11,32,33). The seroprevalence was higher in female animals (4.00%) which may be due to higher stocking of females compared to males, as the male animals are regularly sent for slaughter (for meat). The higher seroprevalence in some pregnant female animals may also be due to the presence of erythritol in the female genital tracts (34). As brucellae lack fructose 6 phosphate kinase and aldolase, they intend to breakdown sugars which primarily use the HMP pathway for energy production such as erythritol and consequently this favours growth of the bacteria (35). More animals were positive in the 1 to 2 years age group which may be due to the sampling nature of the present study. Also, the STAT results in the present study showed more seroprevalence in the organized group (6.67%) over the unorganized group (2.22%) which is not in line with findings of Sharma et al. (31) who reported the unorganized group (3.44 per cent) to be almost similar in seroprevalence to the organized group (3.33 per cent). The reason for higher seroprevalence in an organized set up animals could be close proximity of the animals and also the transmission dynamics and stage of infection. The source of the antigen used for the STAT may also influence its result. In the present study, an in-house produced killed antigen based upon *B. melitensis* was used unlike the *B. abortus* antigens used elsewhere. Also, STAT is usually done for an endemic herd, where the seroprevalence is expected to be consistently high. However, in the current study, only few organized herds were sampled and majority of the samples were from unorganized clusters where farmers maintain few animals (n<10). Also it is to be noted that STAT produces some false positives due to cross reactivity with *Yersinia enterocolitica* ‘O’ Antigen and other shared homophilic lipopolysaccharides (LPS) and heterophilic antigens produced during other febrile illness (36). Thus, the STAT results of the present study point to the scope of taking paired sera samples. Also in chronic diseases like brucellosis, a single most test assay cannot be relied for screening and diagnosis owing to the current recommendation of ‘test and slaughter’ policy, by which valuable germplasm might be lost. Hence, a combination or battery of assays needs to be applied to get plausible calls for declaration of results.

### In total four samples were subjected to 2ME test

One of the samples showed a negative result which implied that the agglutination obtained in STAT for this sample could be due to IgM antibodies indicating an acute infection prior to the class switching of the antibodies. Three out of four samples showed agglutination up to the 1:10 dilutions and this could be due to residual IgG. Thus, in these three samples, the disease may be in the chronic transitional phase wherein both IgG and IgM were present. Thus, STAT and 2ME results in combination thus provide evidence of acute infections and infections which are transitioning into the chronic phase.

**The results of the present study showed seroprevalence by IgG iELISA** fairly higher to some studies (10,32,33) and slightly lower than a study by Kaur et al. (11). Also the higher seroprevalence in female animals and also in organized group of clusters could be explained by the reasons aforementioned in the discussion on the results of STAT. In the current study, the sampling was logically planned and executed (cluster-wise sampling), where it can be explained that organized farms could have more occurrence of brucellosis due to higher density and close proximity of animals. The results also points to the management practices of the organized sector playing a crucial role in the disease dynamics. Also, it is known that iELISA is a highly sensitive test but it is possible for cross reactions to occur with the S-LPS of other bacteria. Hence, it is very important to have a consistent and standard sampling plan while targeting brucellosis by iELISA. The type of iELISA as in targeting only IgG or both IgG and IgM may also influence the results of iELISA. The results of IgG iELISA in the present study reflect the fact persistent infection in the goat populations may be tested for brucellosis using the IgG iELISA and its use as a screening test may be envisaged for caprine brucellosis which may then be confirmed by using other tests such as STAT or 2ME.

### Molecular screening in goat genital swabs

Out of the 120 samples screened, 21 samples (17.50 %), no sample and 6 samples (5.00 %) were positive for the genus specific *16S rRNA* gene for Brucella (Table 3; Fig. 2), species specific *omp31* and *omp2* (Table 3; Fig. 3) genes, respectively by conventional PCR. Also, 48 (40.00 %) samples were positive by TaqMan^®^ real time PCR for *omp31* gene of *B. melitensis* (Table 3). The amplification curve of unknown super-pools in the real time system has been depicted (Fig. 4). The split up of the results of molecular screening in goat genital swabs based on the sex, age group and also, the major clusters were also tabulated previously (Table 2).

**FIG 2.**
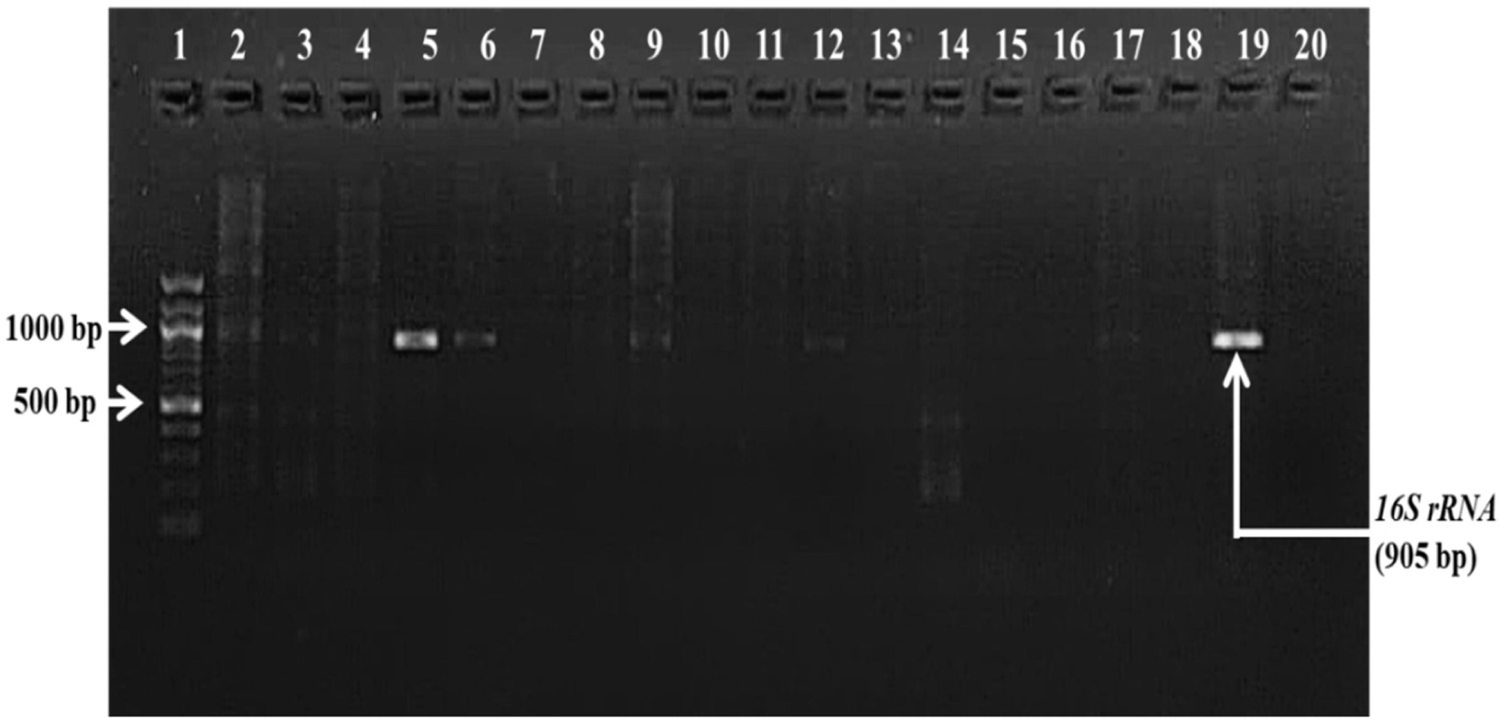
Gel picture of conventional PCR for *16S rRNA* for genus Brucella (Lane 1 – 100 bp DNA marker; Lanes 2, 3, 4, 7, 8, 10, 11, 13, 14, 15, 16, 17, 18 – Negative samples; Lanes 5, 6, 9, 12 – Positive Samples; Lane 19 – Positive control; Lane 20 - Negative control)

**FIG 3.**
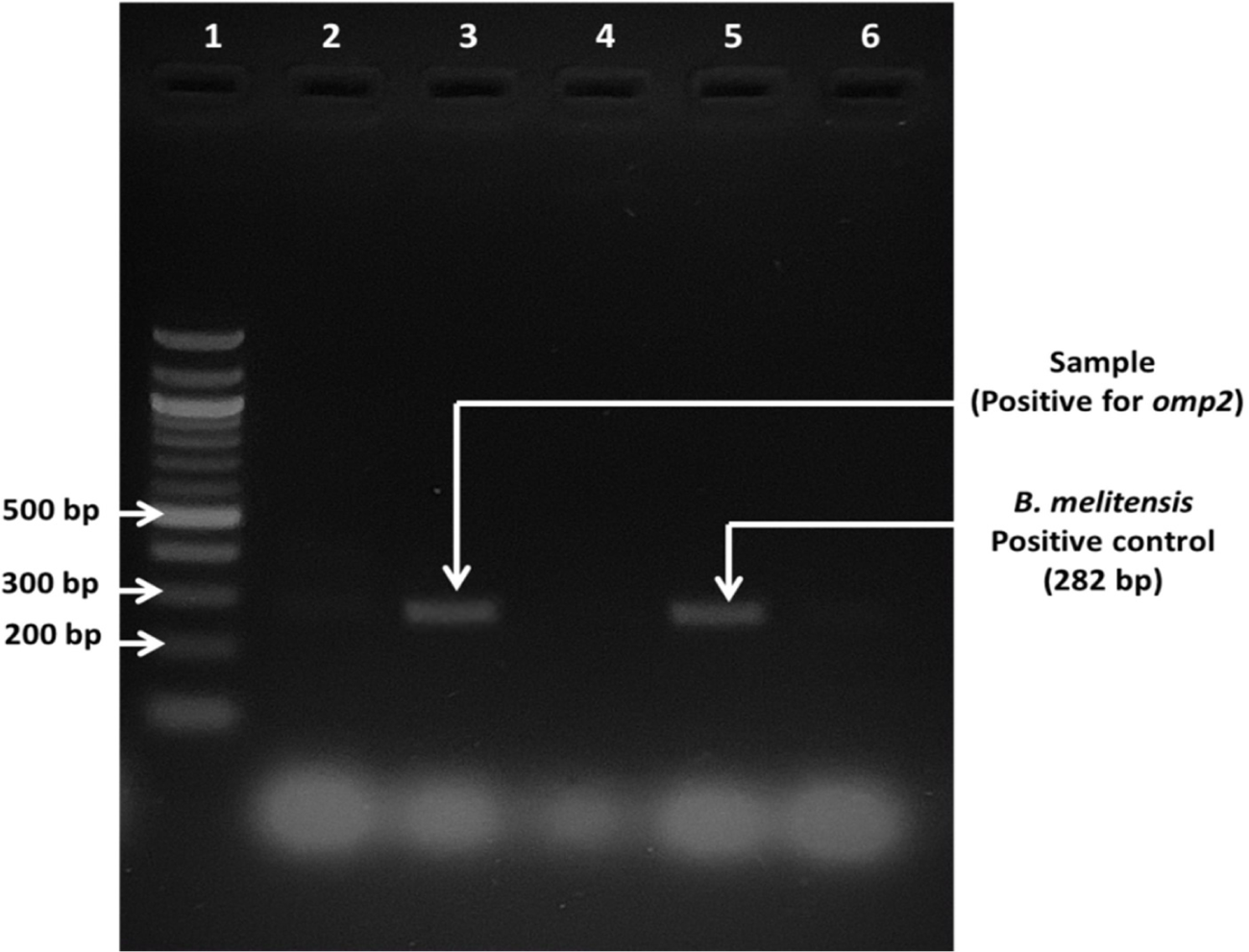
Gel picture of the amplification of *B. melitensis omp2* gene by conventional PCR ( Lane 1-100bp DNA ladder; Lane 2, 4 – Negative samples; Lane 3 – Unknown sample (Positive); Lane 5 – Positive control - *B. melitensis* Biovar 3 ‘Ind’ strain; Lane 6 – Negative control)

**FIG 4.**
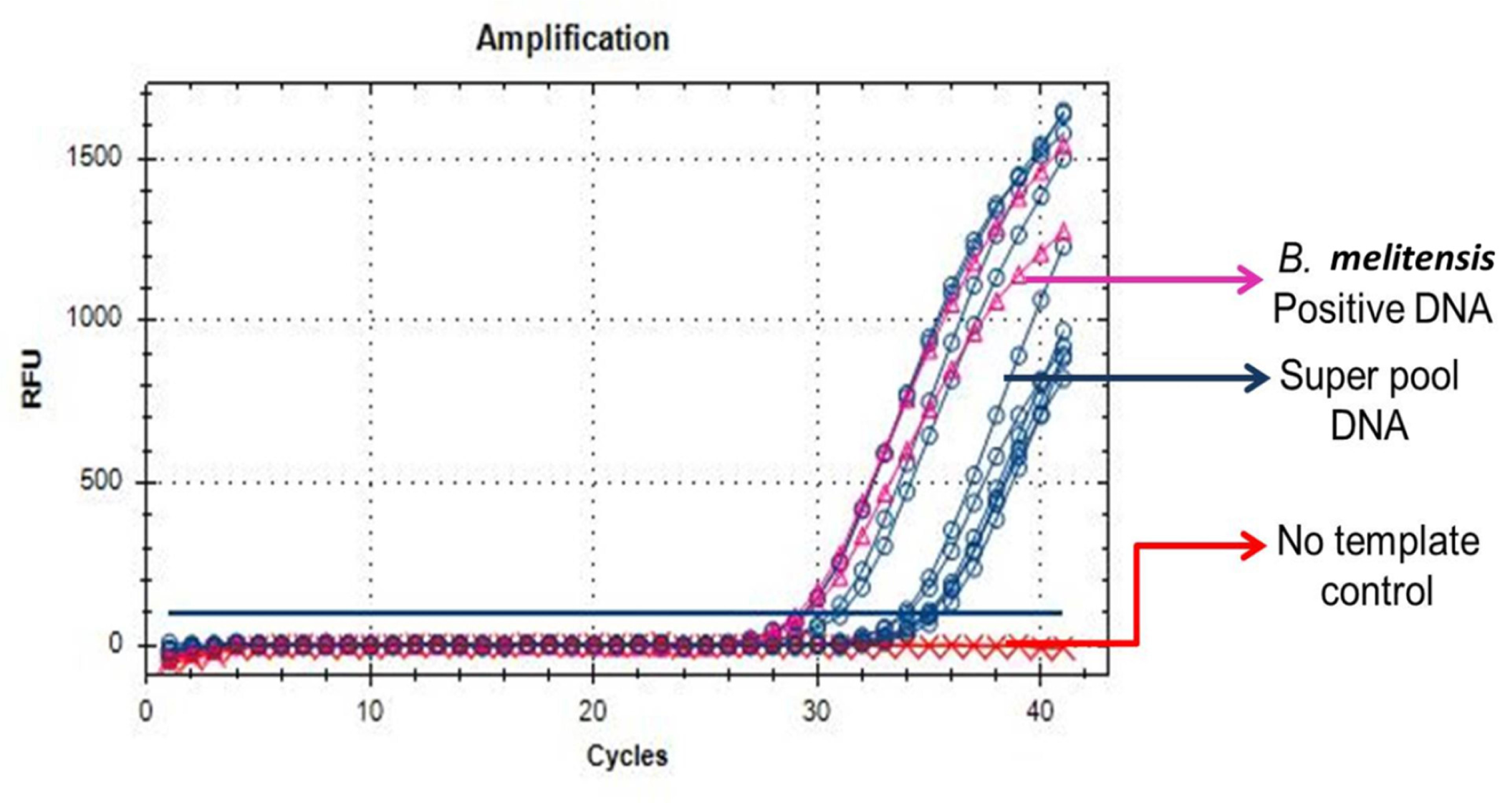
Cq amplification plot of *Brucella melitensis OMP31* TaqMan^®^ Probe real-time PCR for unknown superpools (5) along with positive and negative controls

### Conventional PCR

The detection of genus specific *16S rRNA* gene was in nearly similar order to an earlier study conducted in Mathura, India (16.66%) which included genital swabs (37). The immunological status of the animal with respect to age may be responsible for the susceptibility of the animal to the brucellae. With regard to conventional PCR of the species specific *omp31* and *omp2* genes, *omp31* was not detected in any of the samples but *omp2* was detected in 5.00% of the samples. The reason for missing out on *omp31* may be the low copies of amplicon in the sample and a lower gene base pairs target, which requires a minimum of 10 ng (nanograms) for appreciation of results in EtBr stained gel. Thus for *omp31* gene of *B. melitensis*, the use of nested PCR’s may be explored. However, the *omp2* gene of *B. melitensis* was detected and so it can be targeted for species specific detection of *B. melitensis* from field samples such as genital swabs (38, 39).

### OMP31 TaqMan^®^ qPCR

A study on suspected field samples from goats in U.P., India reported the detection per cent by TaqMan^®^ chemistry to be 49.05 per cent (13) which was higher than the present study. Whereas, a study on breeding bucks in Northern India, did not detect any positives by TaqMan^®^ qPCR in the preputial swab samples (40). Reasons for detection of *omp31* by real time PCR over conventional PCR may be due to the fact that TaqMan^®^ real time PCR is highly specific and sensitive (ability to detect as low as 40 femtograms of the bacterial DNA) (13). Also, specific geographical and climatic conditions which include soil pH, soil type and rainfall and humidity which may influence the survival of the brucellae. Contaminated environment may also reflect on real time PCR owing to its sensitivity (41). Advancing age may be a reason for increased susceptibility to brucellosis. In the present study, the higher occurrence of brucellae in unorganized group of clusters fits an earlier explanation of more occurrence and detection in open grazing systems (42). Also, Another the fact favouring detection is that in an endemic herd there is always an infection cycle maintained in the form of transmission dynamics encompassing various animal, carrion eating birds’ in the vicinity acting as mechanical carriers. However the presence of the organism alone may not be the evidence of disease as confirmation requires concurrence with clinical picture as well as a range of diagnostic techniques.

### Analyses of battery of tests

The analyses of the various diagnostic parameters such as sensitivity, specificity, diagnostic accuracy, positive predictive value (PPV) and negative predictive value (NPV) with respect to *OMP31* TaqMan^®^ qPCR (Table 4) and goodness of fit between various pairs of tests based on Kappa value and McNemar’s Test (Table 5) respectively.

**TABLE 4.** Consolidated results of diagnostic parameters of STAT, IgG iELISA, *16S rRNA* PCR and *omp2* PCR with respect to *OMP31*TaqMan^®^qPCR as standard

| Test | Sensitivity (%) | Specificity (%) | PPV (%) | NPV (%) | Diagnostic Accuracy (%) |
| --- | --- | --- | --- | --- | --- |
| STAT | 6.25 | 98.61 | 75.00 | 61.21 | 61.67 |
| IgG iELISA | 14.58 | 79.17 | 31.82 | 58.16 | 53.00 |
| *16S rRNA* PCR | 41.67 | 98.61 | 95.24 | 71.72 | 75.83 |
| *omp2* PCR | 12.50 | 100 | 100 | 63.18 | 65.00 |

PPV – Positive Predictive Value
NPV – Negative Predictive Value

**TABLE 5.** Comparison of agreement of diagnostic tests for caprine brucellosis based upon Kappa value (for agreement) and McNemar’s test (for *p* value)

| Diagnostic tests compared | Kappa value | 95 % Confidence Interval | *p* value | Strength of agreement |
| --- | --- | --- | --- | --- |
| STAT vs *OMP31* TaqMan<sup>®</sup> qPCR | 0.057 | -0.029 to 0.144 | 0.0001 | Slight agreement |
| IgG iELISA vs *OMP31* TaqMan<sup>®</sup> qPCR | -0.069 | -0.219 to 0.082 | 0.0008 | No agreement |
| *16S rRNA* PCR vs *OMP31*TaqMan<sup>®</sup> qPCR | 0.444 | 0.294 to 0.594 | 0.0001 | **Moderate agreement** |
| *omp2* PCR vs *OMP31* TaqMan<sup>®</sup> qPCR | 0.146 | 0.038 to 0.255 | 0.0001 | Slight agreement |
| STAT vs PCR (*omp2*) | 0.167 | -0.173 to 0.506 | 0.7237 | Slight agreement |
| IgG iELISA vs PCR (*omp2*) | -0.008 | -0.143 to 0.128 | 0.0033 | No agreement |
| *16S rRNA* PCR vs PCR (*omp2*) | 0.398 | 0.170 to 0.625 | 0.0003 | **Fair agreement** |
| STAT vs IgG iELISA | 0.022 | -0.115 to 0.158 | 0.0005 | Slight agreement |
| STAT vs *16S rRNA* PCR | 0.110 | -0.128 to 0.258 | 0.0005 | Slight agreement |
| IgG iELISA vs *16S rRNA* PCR | 0.065 | -0.128 to 0.158 | 1.000 | Slight agreement |

**Kappa value** (< 0: No agreement; 0.00 and 0.20: Slight; 0.21 and 0.40: Fair; 0.41 and 0.60: Moderate; 0.61 and 0.80: Substantial; 0.81 and 1.00: Almost perfect)

For any epidemiological setting, one standard test may not be conclusive and so using a battery of tests is recommended for diagnosis of brucellosis (19). Also, a combination of tests (both direct and indirect) for confirming the disease is recommended over reliance on a single test (43). Considering the above aspects, it was also suggested that evaluation of diagnostic tests was required on a study-to-study basis as no universal standard is available for either direct or indirect diagnosis of caprine brucellosis caused by *B. melitensis* because of a number of variables in each study (43). Hence, serological tests were compared with the molecular tests in the present study to show the extent of difference in their diagnostic abilities. It is not rational to compare the serological with molecular, but often diagnosis is made solely on serological tests for brucellosis which is misleading and cause poor decisions. Again, higher sensitivity with low specificity (iELISA etc.) is reason for over-reporting of brucellosis. Consolidated results of diagnostic parameters of STAT, IgG iELISA, *16S rRNA* PCR and *omp2* PCR with respect to *OMP31*TaqMan® qPCR as standard show that with respect to the serological tests, the IgG iELISA was more sensitive (14.58%) compared to STAT (6.25%). However the STAT showed more specificity and diagnostic accuracy compared to IgG iELISA (Table 4). This points to the higher sensitivity of iELISA in detecting antibodies and that its combination with STAT for diagnosis of disease may be used for obtaining fairly higher specificity notwithstanding the fact that only slight agreement may be obtained in the results (Table 5). With respect to the molecular tests, the *omp2* PCR showed 100% specificity and positive predictive value while the *16S rRNA* PCR had higher diagnostic accuracy. This shows the fact that qPCR is more sensitive to other conventional PCRs as reported earlier (13). Also, while targeting species specific genes for *B. melitensis*, a combination of *omp2* PCR with qPCR may be explored to obtain higher specificity and sensitivity. Also, the agreement among the various tests based upon the Kappa value and McNemar’s test has been shown which reaffirm the fact the findings stated above (Table 5).

### PCR-RFLP

The suggestive results of RFLP of the *omp2* gene product revealed the strain of *B. melitensis* in Puducherry to be Biotype/Biovar 3 which is a first of its kind report from Puducherry region (Fig. 5).

**FIG 5.**
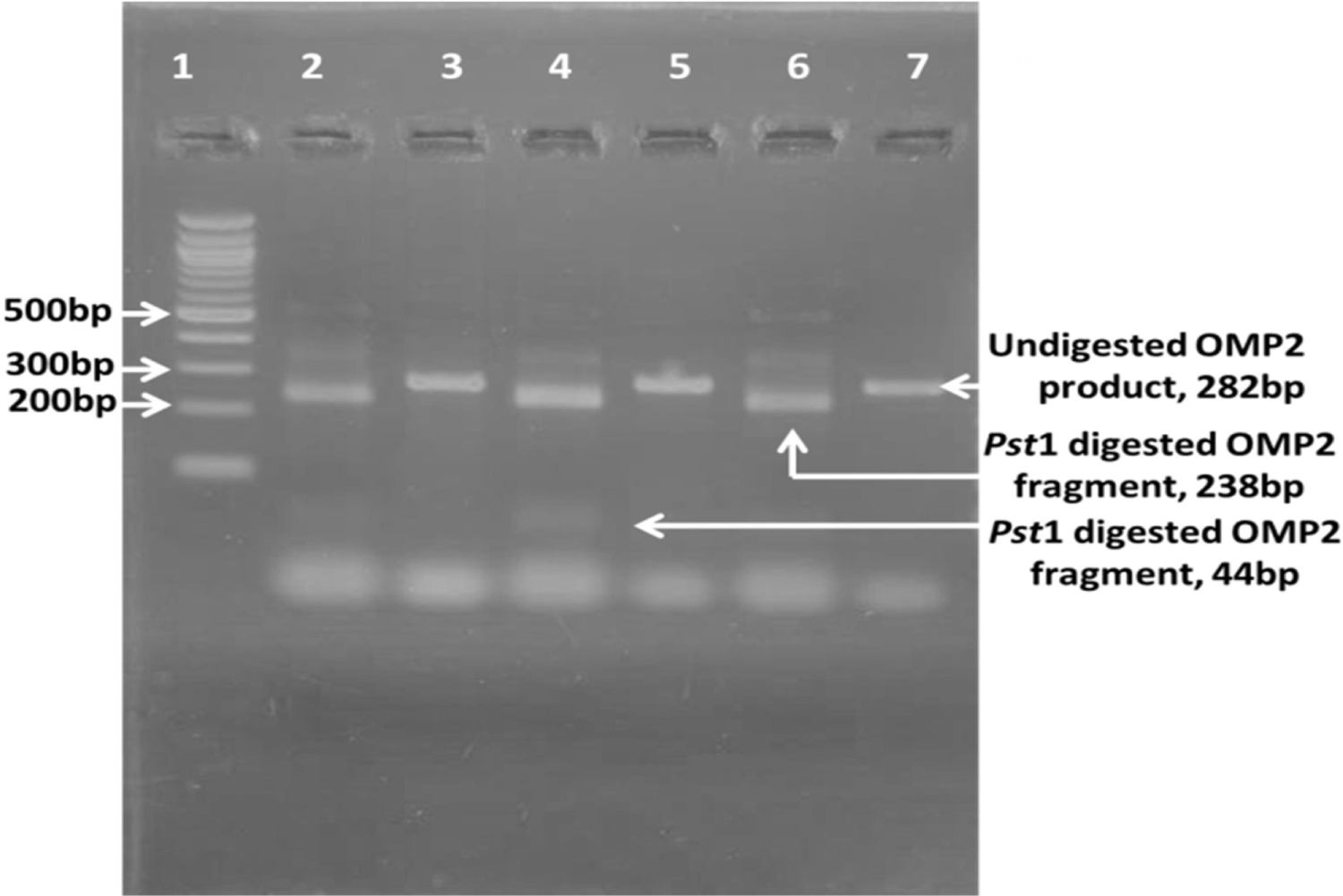
Gel picture showing PCR-RFLP of *omp2* gene of *Brucella melitensis*. Lane 1-100bp DNA ladder; 2,4 & 6 –*Pst*I digested *omp2* product (238bp and 44bp fragments) suggestive of *B. melitensis* Biovar 3; 3, 5 & 7-Undigested *omp2* gene amplicon (282bp) of *Brucella melitensis*

Prior study in Mathura India, reported similar findings wherein *B. melitensis* Biovar 3 was the most prevalent in India (12) and it is the most predominant of all the biovars reported otherwise (44). However, it is to be noted that other workers have reported Biotype 1 strain of *B. melitensis* from Bengaluru, India, based on PCR – RFLP analysis of the *omp2* gene product (14). The use of PCR – RFLP for the *omp2* gene product is a fairly quick method for strain analysis on suspected field samples for *B. melitensis* (12). Thus, for field isolates of *B. melitensis*, The RFLP of *omp2* gene product is a reliable tool for strain identification of *B. melitensis*.

### Serological tests on human blood serum

Out of 30 samples tested from human subjects, while no sample was positive by RBT or STAT, 10 samples (33.33 %) were positive by IgG iELISA. The results of serological tests in human subjects based on the broad groups, gender and age of the subjects have also been accordingly tabulated (Table 6).

**TABLE 6.** Tabulation of the results of serological tests on human subjects for brucellosis in Puducherry

| Category of human subjects | T | RBT |  | STAT |  | IgG iELISA |  |
| --- | --- | --- | --- | --- | --- | --- | --- |
|  |  | P | % | P | % | P | % |
| *Group wise* |  |  |  |  |  |  |  |
| Interns/Post Graduate students | 15 | 0 | 0.00 | 0 | 0.00 | 3 | 20.00 |
| Farmers | 15 | 0 | 0.00 | 0 | 0.00 | 7 | 46.67 |
| *Gender wise* |  |  |  |  |  |  |  |
| Male | 18 | 0 | 0.00 | 0 | 0.00 | 5 | 27.78 |
| Female | 12 | 0 | 0.00 | 0 | 0.00 | 5 | 41.67 |
| *Age wise* |  |  |  |  |  |  |  |
| Up to 25 years | 12 | 0 | 0.00 | 0 | 0.00 | 1 | 8.33 |
| 25 to 50 years | 17 | 0 | 0.00 | 0 | 0.00 | 8 | 47.06 |
| Above 50 years | 1 | 0 | 0.00 | 0 | 0.00 | 1 | 100 |
| Total | 30 | 0 | 0.00 | 0 | 0.00 | 10 | 33.33 |

T-Number tested
P-Number positive

**The inability of the RBT to detect positivity in human subjects** in the present case may be due to the targeted heterogeneous population. Lower titres of antibody in case of a waning or chronic infection may not show macroscopic agglutination by a coloured antigen test alone and hence require confirmation by other tests (2). The nature of the antigen used in the test may also play a significant role in detecting brucellosis in human subjects. Another drawback with using the RBT is its cross reactions with other S-LPS of bacteria such as *Yersinia enterocolitica* O:9, *Escherichia coli* O:157, *Vibrio cholera*, *Salmonella* O:30 and others (2). Hence the RBT in case of human subjects for the use of screening brucellosis may be used as a preliminary test but it is prone to drawbacks and hence reliance on RBT alone is not recommended for diagnosis of brucellosis in humans risk groups.

The results of the present study resembled reports on STAT in a study conducted in Gujarat (45) and a study carried out earlier in Puducherry (46). On the other hand, studies in Kerala and Jammu found the seroprevalence in human subjects to exhibit a range from 1.60 % to 9.09 % by STAT (15, 16). The source of the plain antigen may be a cause for the results of the STAT i.e. the present study used a *Brucella melitensis* plain antigen in contrast to *B. abortus* plain antigen used elsewhere. Also, insufficient titre of antibodies in the case of chronic or waning infections may result in absence of macroscopic agglutination. Another reason for the inability of STAT in the present study to show positives may be because of prozone phenomenon and hence pre-treating the serum with heat may be explored in certain cases (2). While cross reactions have a chance of occurring in STAT even in the case of human subjects; the use of STAT alone as a confirmatory diagnostic test may be contraindicated in diagnosis of human brucellosis owing to its shortcomings.

### In the present study, seroprevalence by IgG iELISA in human subjects was 33.33%

In contrast, a study conducted earlier in Puducherry by Ranganathan et al. (46) did not show any positives. Whereas, a study on human patients with PUO in Goa, India showed lower seroprevalence of 4.96% by IgG iELISA (47). A study in Karnataka, India on 1050 human sera samples from risk groups showed lower seroprevalence at 6.76% (48). Another study by Padher et al. (45) in central Gujarat also showed lower seroprevalence by IgG iELISA at 12.00%. The detection by IgG iELISA alone may not give a comprehensive detail of the disease in human subjects until it is co-related with results of other tests, clinical signs, co-morbidities, risk factors, etc. (48). Lower titres of antibodies in the case of waning infections and possibility of previous exposure to the antigens may be detected by IgG iELISA. Also, in chronic cases and relapse of the disease in human subjects, the use of IgG iELISA may be of value as previously described (2).

Higher seroprevalence of brucellosis human risk groups such as veterinary students reinstates the need to follow safe handling practices Serological evidence by IgG iELISA shows probability of previous exposure and also provides considerable insight into circulaltion of the bacteria among both unorganized and organized sectors. The higher seroprevalence in farmers group may reflect the higher prevalence of the organism in the field conditions and hence higher risk in the farming communities to whom appropriate education and hygienic handling techniques need to be explained by prospective intervention studies.

## CONCLUSION

The seroprevalence of caprine brucellosis in Puducherry, India by RBT, STAT and IgG iELISA was zero, 3.33% and 18.33% respectively in goats. With regard to RBT, the need to develop *B. melitensis* specific coloured antigens is required. The IgG iELISA may be used as a screening test for caprine brucellosis in larger goat populations owing to its better sensitivity and confirmation may be carried out by other tests such as STAT and 2ME. The higher seroprevalence by IgG iELISA in goats indirectly suggests that large number of animals in the geographical areas sampled could be continuously exposed to the pathogen. Population screening also emphasises the need to evaluate the on-going control programmes in a politico-geographical entity for devising better control methodologies in the future. However, for detecting the current status of Brucella infection, IgM iELISA should be used. Molecular screening revealed occurrence of genus specific *16S rRNA* gene in 17.50% and species specific *omp2* gene in 5.00% of the genital washings by conventional PCR. The *OMP31*TaqMan® qPCR is advocated for the molecular detection of *B. melitensis* as it detected 40 % positivity (which included all samples positive in conventional PCR) in the genital swab samples. The optimization and use of a battery of tests is projected to be of value on knowing the status of caprine brucellosis. Also, in future, steps are required to differentiate the live brucellae from dead bacilli using mRNA based PCR detection methods which precisely could improve the understanding of the passive and active carrier status. By this, it is possible to accurately report and adjust the brucella prevalence as serological tests and other tests with higher sensitivity tend to over-report the prevalence of brucellosis. In a first, the strain of *B. melitensis* in Puducherry is reported to be Biovar 3 based on the suggestive result of the RFLP of the *omp2* gene product. Serological and molecular evidence of caprine brucellosis establishes the need for regular screening, monitoring of movement, surveillance and reporting of disease in goats. Also, vaccination in goats may be explored. Human risk groups need to be screened for brucellosis on a regular basis as there is evidence of seroprevalence by IgG iELISA (33.33 %) indicating indirectly the exposure to the pathogen. Also, in human handlers such as farmers, intervention studies to train them on safe management and handling practices need to be carried out. The present study apart from being cross-sectional, serves as a base warranting and providing the scope for detailed spatial and temporal studies to be carried out in future regarding caprine brucellosis and its public health significance. Knowing the public health implications of brucellosis due to *B. melitensis*, the multi-sectorial “One Health Approach” to frame diagnostic, control and eradication strategies needs to be explored given the always evolving disease dynamics and epidemiology.

## Data Availability

All data produced in the present work are contained in the manuscript

## Acknowledgements

The research was carried out under a memorandum of understanding between Rajiv Gandhi Institute of Veterinary Education and Research (RIVER), Puducherry, India and Indian Council for Agricultural Research – Central Institute for Research on Goats (ICAR – CIRG), Uttar Pradesh, India. The authors acknowledge the grants and infrastructure aid provided by the participating institutions for this Master’s programme research work.

## Competing Interests/Conflict of Interest

The authors do not have any conflict of interests.

## Notes

### Competing Interest Statement

The authors have declared no competing interest.

### Author Declarations

Institute Ethical committee (IEC), Indira Gandhi government medical College and hospital, Kadirgamam, Pondicherry

